# Feasibility and acceptability of hepatitis C virus self-testing models among high-risk groups in Nasarawa, Nigeria; exploratory cross-sectional analysis of an implementation study

**DOI:** 10.1101/2025.11.17.25340458

**Authors:** Victor Abiola Adepoju, Chidinma Umebido, Kristina Grabbe, Molly Strachan, Catharine Laube, Oluwafunke Odunlade, Ganiyu Jamiyu, Bashorun Adebobola, Ibrahim Adamu Alhassan, Adetiloye Oniyire, Cheryl Johnson, Karin Hatzold, Yasmin Dunkley

**Author notes:** These authors contributed equally to this work (Victor Abiola Adepoju, Chidinma Umebido, Kristina Grabbe, Molly Strachan, Yasmin Dunkley). These authors also contributed equally to this work (Oluwafunke Odunlade, Ganiyu Jamiyu).

## Abstract

**Background:** Hepatitis C virus (HCV) infection remains a major public health challenge, with significant gaps in diagnosis and treatment in resource-limited settings. Hepatitis C self-testing (HCVST) offers a potential strategy to expand access, particularly in HIV clinical settings. We evaluated feasibility and acceptability of HCVST among high-risk populations in Nasarawa State, Nigeria when provided at antiretroviral (ART) clinics and one-stop shops (OSS) serving key populations (KP).

**Methods:** 2,000 participants were enrolled between May 2023 and December 2023. Participants tested with either blood-based or oral fluid HCVST, and with or without health worker support. Follow-up documented results and linkage to qualitative RNA PCR testing and treatment. Feasibility was assessed across the HCV care cascade using chi-square tests. Acceptability was evaluated using a post-test survey score derived through factor analysis; associations between low acceptability (lowest 10%) and participant characteristics were explored through regression analyses. Outcomes were compared by facility type (ART clinic vs. OSS). Free-text responses were thematically analyzed to contextualize findings.

**Results:** HCVST was feasible and acceptable. Of 226 reactive HCVST results (11.3%), 99.1% received RNA PCR testing. Among those with detectable RNA, 92% initiated treatment and 97% completed therapy. However, differences were observed by facility type. Participants in ART clinics were older, more likely to be female, and showed higher reactivity (15% vs. 8%) and treatment uptake (96% vs. 83%) than OSS clients. Acceptability was higher in ART clinics than OSS.

**Conclusions:** HCVST was both feasible and acceptable in Nasarawa State, with some observed variations by facility type. These findings demonstrate that with differentiated service delivery models and adequate support for linkage, HCVST can increase HCV diagnosis, linkage to care, and treatment among high-risk groups in Nigeria, supporting integration of HCVST into national viral hepatitis elimination strategies.

## Introduction

Globally, an estimated 50 million people live with chronic hepatitis C virus (HCV) infection, and about 240,000 die each year from complications such as chronic liver disease, cirrhosis, and cancer [1]. While no vaccine exists, direct-acting antivirals (DAAs) now cure approximately 95% of cases [1–3], driving global momentum toward elimination. Yet major service gaps exist, particularly in low- and middle-income countries (LMIC). In 2022, only 36% of people with HCV globally knew their status, and just 20% of those diagnosed had initiated treatment [1,4].

HCV disproportionately affects vulnerable populations, especially people living with HIV (PLHIV) and key populations (KP), including people who inject drugs (PWID), men who have sex with men (MSM), and sex workers (SW) [5]. These groups face overlapping HCV and HIV risk factors such as needle sharing and condomless sex, compounded by stigma, discrimination, and legal barriers that limit care. WHO estimates 2.3 million or 5.9% of PLHIV have current or past HCV infection [6,7]. Sub-Saharan Africa bears the second-largest burden of HIV-HCV co-infection with around 430,000 cases [7]. Globally, more than half of co-infections occur among PLHIV who inject drugs [7]. PLHIV who are sex workers have 1.4-6.8 times higher odds of also living with HCV, while MSM and PWID face 4-13 times higher odds [7]. Co-infection accelerates liver disease progression, contributing significantly to morbidity and mortality [8].

Nigeria bears one of the highest HCV burdens, with approximately 2.4 million people chronically infected (1.1% prevalence) [9]. Prevalence peaks at ages 50-54 years (3.3%) and is lowest among 15-19-year-olds (0.4%) [9]. Among PLHIV, HCV prevalence is estimated at 1.1% [9], but some studies suggest rates up to 2.3% [10]. Data for key populations are limited but overlapping risks with PLHIV make them a priority for testing and treatment. Awareness is critically low, over 80% of Nigerians with viral hepatitis do not know their status [11].

Nasarawa State carries a particularly high HCV burden, with an estimated 13.2% prevalence [9]—more than 250,000 people. In 2020 the state launched a five-year micro-elimination plan focusing on PLHIV and KP, but progress has been limited. By study initiation in 2023, only 85,000 people had been screened and 1,500 treated, far below the goal of testing 2.4 million and treating 124,000 by 2025 [12]. Expansion of HCV services has been constrained by high costs, stigma, and low public awareness. While HIV antiretroviral therapy (ART) clinics offer some capacity, community-based one-stop shops (OSS) for KP often lack HCV testing, limiting access for those most at risk.

HCV self-testing (HCVST) has emerged as a promising innovation to address these barriers. Endorsed by WHO in 2021, self-testing methods can expand access to HCV screening and enable higher rates of HCV treatment and cure among underserved populations [13]. Multi-country studies, including in Kenya and Egypt, found HCVST feasible and acceptable, with most users able to perform tests independently and willing to recommend them [14–19]. Benefits cited in those studies include privacy, convenience, and autonomy, while challenges included the need for confirmatory testing, linkage to care, and support to mitigate misuse or distress [20]. Blood-based and oral fluid-based kits are available but not yet widely used in Nigeria, constrained by costs, regulatory barriers, and limited country-level evidence.

As Nigeria advances toward HCV elimination, especially in high-prevalence states like Nasarawa, context-specific evidence on HCVST is essential. Understanding feasibility and acceptability among PLHIV and KP can inform policy, drive down costs, and support scale-up. In particular, assessing HCVST in decentralized, non-clinical sites is critical, given gaps in current service delivery. This study examined the feasibility and acceptability of HCVST in ART clinics and OSS in Nasarawa State, Nigeria to guide future introduction and scale up.

## Methods

### Study design

This cross-sectional analysis of an implementation study explored feasibility and acceptability of HCVST among persons at high risk for acquiring HCV in Nasarawa State in Nigeria from May 30, 2023 to April 30, 2024 with recruitment conducted between May 30, 2023 to December 4, 2023. HCV care cascade outcomes were prospectively tracked throughout the implementation period. This manuscript follows the Strengthening the Reporting of Observational Studies in Epidemiology (STROBE) guidelines [21]. Additional exploratory qualitative analysis was conducted on free-text responses to enrich acceptability analyses.

### Setting

The study was conducted in Nasarawa State, selected due to the high prevalence of both HCV and HIV and the state’s commitment to addressing HCV as evidenced by efforts outlined in the HCV micro-elimination plan. Four sites were identified for participation in the study: two ART clinics serving PLHIV in public health facilities, and two community-based OSS serving KP. Sites were purposively selected based on populations served, level of care provided, and the presence of systems to provide follow-up services including linkage to confirmatory testing and HCV treatment.

### Study participants

The study enrolled 2,000 people aged 18 years and above who were living with HIV and/or met the study definition of KP and did not know their HCV status. Study populations were selected due to reported risk factors for HIV and HCV co-infection, specific risk behaviors that make them more susceptible to acquiring HCV, and experiencing barriers to accessing conventional health care services, including stigma, discrimination, and legal and social barriers.

### Implementation procedures

Clinicians in study sites were given a script to read to existing clients during routine clinic visits, health talks, or through routine community outreach. Clients interested in learning more about the study were introduced to a trained research assistant (RA) who described the study in detail. Once enrolled, study participants completed a received education about HCV transmission, prevention, and treatment availability, as well as instructions on how to conduct either blood-based or oral fluid-based HCVST. Professional HCV RDTs were available for all clients who were interested in HCV testing but did not consent to participate in the study.

Participants could choose either a blood-based lateral flow test (First Response® HCV Card Test (Self-Test), Premier Medical Corps, Ltd., Gujarat, India) or an oral fluid-based rapid test (OraQuick® HCV Self-Test, OraSure Technologies, Inc., Pennsylvania, USA). Once a test type was exhausted, participants could still enroll in the study but no longer had the option of choosing their test type. Participants also had the option of conducting HCVST on-site in the presence of a health worker (observed), or on- or off-site without a health worker (unobserved).

Those who chose unobserved HCVST could access on-demand assistance if they encountered difficulties, and were contacted by phone, text, WhatsApp, or home visit two days after receiving the HCVST to report their test results and support linkage to follow up services as necessary. A post-test survey to assess user experience was administered to all study participants.

Participants with a reactive HCVST were linked with a nearby health facility for an HCV RNA PCR test to confirm the presence of HCV and inform a treatment plan, as well as a liver function test, and clinical evaluation. Participants received up to five phone calls, messages, or home visits to support this linkage (days 3, 7, 14, 21, and 28). Those with detectable HCV RNA received treatment with DAAs (sofosbuvir/velpatasvir) per national guidelines, which was provided free of charge to all study participants. Participants were followed up by phone throughout the course of treatment to confirm completion of DAAs and document cure as defined by the results of viral load testing done approximately 12 weeks post treatment. HCVST reactive study participants who also reported injecting drugs were offered the opportunity to engage their sexual or injecting drug use partners in the study—they were given a WhatsApp/SMS number to connect with study staff, who provided support and information about the study and HCV to the partners or referred them to a nearby HCV center for conventional testing and treatment if they preferred. Partners interested in participating in the study could pick up an HCVST from the study site, or directly from the initial study participant (secondary distribution).

### Exposure variables

Demographic, behavioral and clinical characteristics of study participants, as well as HCVST type used (blood-based or oral fluid), approach (observed or unobserved), and facility type (ART clinic or OSS) were collected on an intake form administered upon study enrollment (**S1**) and from an endline survey completed post-test (**S2**).

Demographic variables collected were self-reported, grouped into: gender (male/female), age (18-24, 25-34, 35-44, 45-54 and 55+) education (no formal education, primary, high school, tertiary), occupational status (employed for wages, self-employed, out of work, student or “other”), marital status (single, never married, married/in domestic partnership, widowed, divorced/separated). Data elements chosen aligned with national program data capture and retained sufficient statistical stability within groups.

Exposure variables hypothesized as associated with feasibility and acceptability were facility type (ART clinic or OSS), participant HIV status (positive, negative or unknown) and KP identity. KP identity was reporting any one of the following characteristics: persons who inject drugs (PWID—reported ever injected drugs for non-medical use), men having sex with men (MSM—reported male gender and same sex activity), sex worker (reported ever received goods or services for sex), and partners of sex workers (people who had ever paid for sex). All variables were identified through self-report, with HIV status confirmed through clinical records where possible.

### Outcome variables

#### Feasibility

Feasibility was explored through study uptake and care cascades; uptake of HCVST (study enrollment, use of HCVST), results (reactive, non-reactive), and—if reactive—access to follow up services (RNA PCR testing, treatment uptake, treatment completion and viral suppression) across all four sites. A study logbook (**S3**) tracked these follow-up service data as self-reported by participants and later verified with their HCV treatment center. Client data from the intake form was linked with that in the logbook using a unique identifier.

#### Acceptability

Acceptability was explored through a composite client acceptability score constructed using exploratory and confirmatory factor analyses (EFA and CFA) derived from items from the endline satisfaction survey administered post-test which included structured Likert responses and free-text items. Data was entered in-person for those who completed the HCVST on-site, and by phone for those who used the test off-site.

Factor analysis was selected as the most appropriate method for the acceptability score given the latent, multidimensional nature of psychological acceptability constructs [22–23]. Seventeen indicators were identified as conceptually related to acceptability of HCV self-testing, drawing from four hypothesized domains: (i) ease of use, (ii) confidence and confidentiality, (iii) self-efficacy in managing care, and (iv) future use intentions. Items unrelated to the construct—such as HCV knowledge or willingness to pay—were excluded. All indicators used 5-point Likert response options, coded such that higher values reflected greater acceptability. Complete case analysis was used (N = 1868), given minimal item-level missingness.

Internal consistency for the full item pool was high (Cronbach’s α = 0.91), with sampling adequacy confirmed via the Kaiser-Meyer-Olkin test (KMO = 0.92) and Bartlett’s Test of Sphericity (p < 0.001), supporting factorability. EFA revealed a three-factor solution that aligned with initial conceptual domains (empowerment (5 items), ease of use (6 items), and future use intentions (3 items) except for the ‘confidence and confidentiality’ items, which showed low loadings and were excluded (3 items)).

A 3-factor CFA was conducted on the remaining 14 indicators using the WLSMV estimator to account for ordinal-level Likert data. Model fit was good (CFI = 0.996; TLI = 0.995, RMSEA = 0.060; SRMR = 0.042). A second-order CFA specifying an overarching acceptability construct as a latent factor that explains shared variance across three first-order domains retained similarly strong fit indices (CFI = 0.996; TLI = 0.995; RMSEA = 0.060; SRMR = 0.042), supporting the validity of a single composite acceptability score. Participant-level factor scores were generated from the final model using regression (least squares). Given generally high rates of acceptability and left skew data distribution, the continuous score was split into a binary outcome between the lowest 10% of acceptability scores and the remaining acceptability scores for statistical analysis (**S4**).

### Statistical analysis

Data analysis was conducted in R studio and Excel for qualitative analysis.

Participant throughput was described; characteristics were described and compared by facility type using Pearson’s chi-square test with missingness described. Feasibility was explored through study enrollment, test results and linkage across the clinical care cascade, alongside differences by facility type (ART clinic vs. OSS), HIV status (people living with vs. without HIV) and KP identity (versus no reported KP identity). Due to very high collinearity between facility type, HIV status, and KP identity (almost all ART participants were PLHIV and non-KP; nearly all OSS participants were KP), facility type was retained as the sole exposure.

Acceptability was described as a continuous outcome and defined as a binary outcome (lowest 10% of acceptability vs. acceptable). Continuous acceptability scores were explored visually using histogram and boxplots by age, gender, educations status and facility type, with proportions in the lowest 10% of acceptability identified. We modelled binary acceptability by facility type using logistic regression, adjusting for age, gender, and education status as a priori confounders. Qualitative thematic analysis was used to provide explanatory insights into quantitative cascade differences and acceptability. Free-text responses were examined by site type (ART clinic vs. OSS) to identify contextual differences in provider and participant accounts that could plausibly explain variations in cascade outcomes and acceptability.

### Ethics

Ethical approval for the study was obtained from the John Hopkins University Bloomberg School of Public Health Institutional Review Board (BPSH #20755), National Health Research and Ethics Committee in Nigeria (approval number NHREC/01/01/2007-17/07/2022), Nasarawa State Ministry of Health Ethics Review Committee (approval number NHREC 18/06/2017), and World Health Organization Ethics Review Committee (WHO ERC 3809). Study participants provided written informed consent.

## Results

### Participant Throughput

A total of 2,000 people were enrolled and consented to participate in the study. Five participants took HCVST kits home but did not return to complete the endline survey, therefore no demographic or clinical information was recorded for those participants, and they were excluded from the analysis. The final analysis included data from 1,995 participants. 11 of these participants reported reactive HCVST results but did not complete the endline survey during the study period; their information is included and missingness described (**Figure 1**).

**Figure 1.**
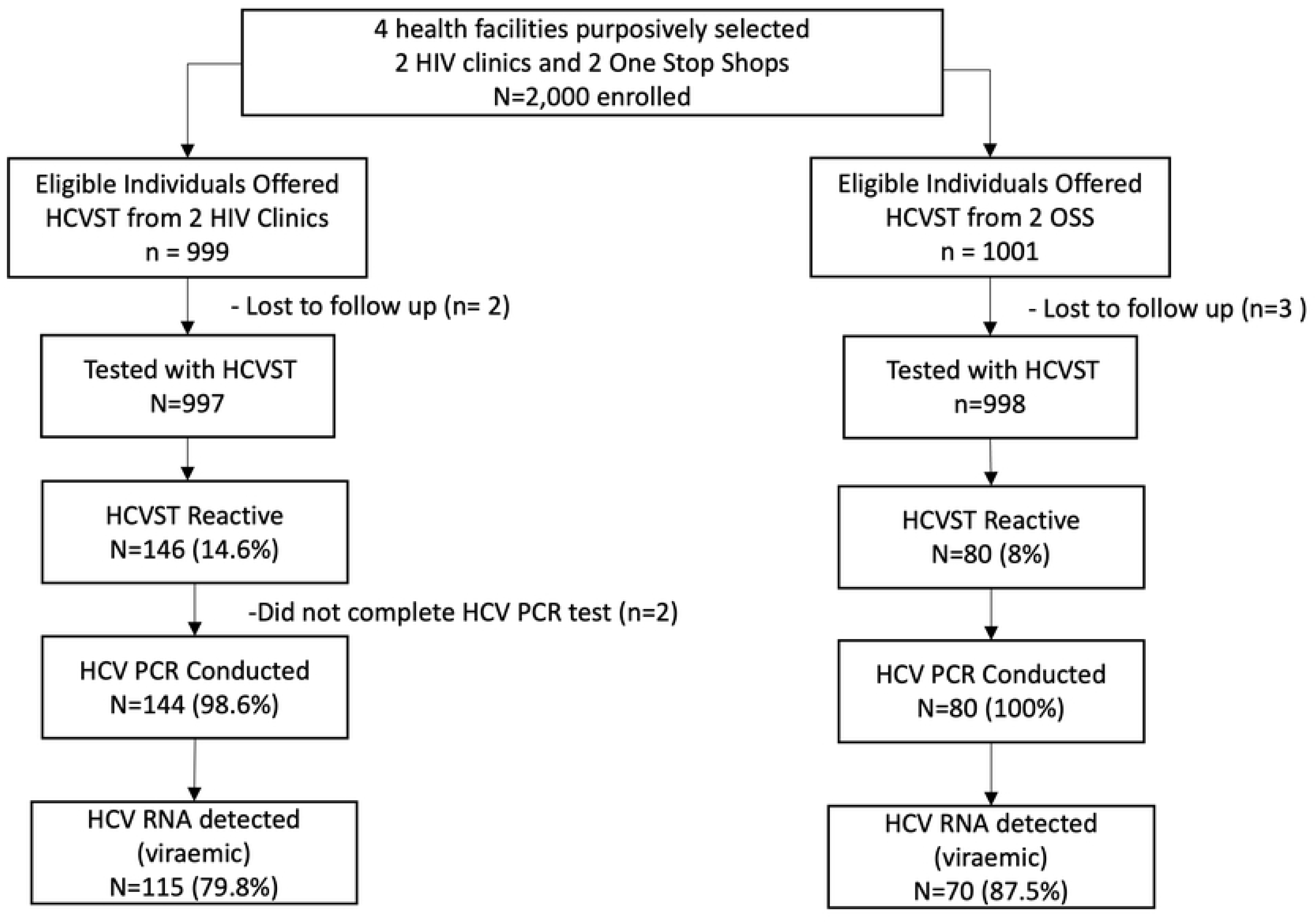
Study Throughput.

### Participant Characteristics

#### Sociodemographic and behavioral factors

Study participants were mostly female (60%), 25-44 years old (65%), and the majority had at least high school education (66%), were currently employed (66%), were not currently married or in a domestic partnership (53%), did not report KP identity characteristics (51%) and were living with HIV (65%). Of the 978 individuals reporting KP identity characteristics, only 16 (2%) were from an ART clinic, while 962 (98%) were from an OSS (Table 1). As a proportion of all OSS attendees (where participants could report multiple KP identities, 314 (32%) identified as MSM, 458 (46%) as sex workers, 279 (28%) as PWID, and 124 (12%) as partners of sex workers (**Table 1**).

**Table 1.**
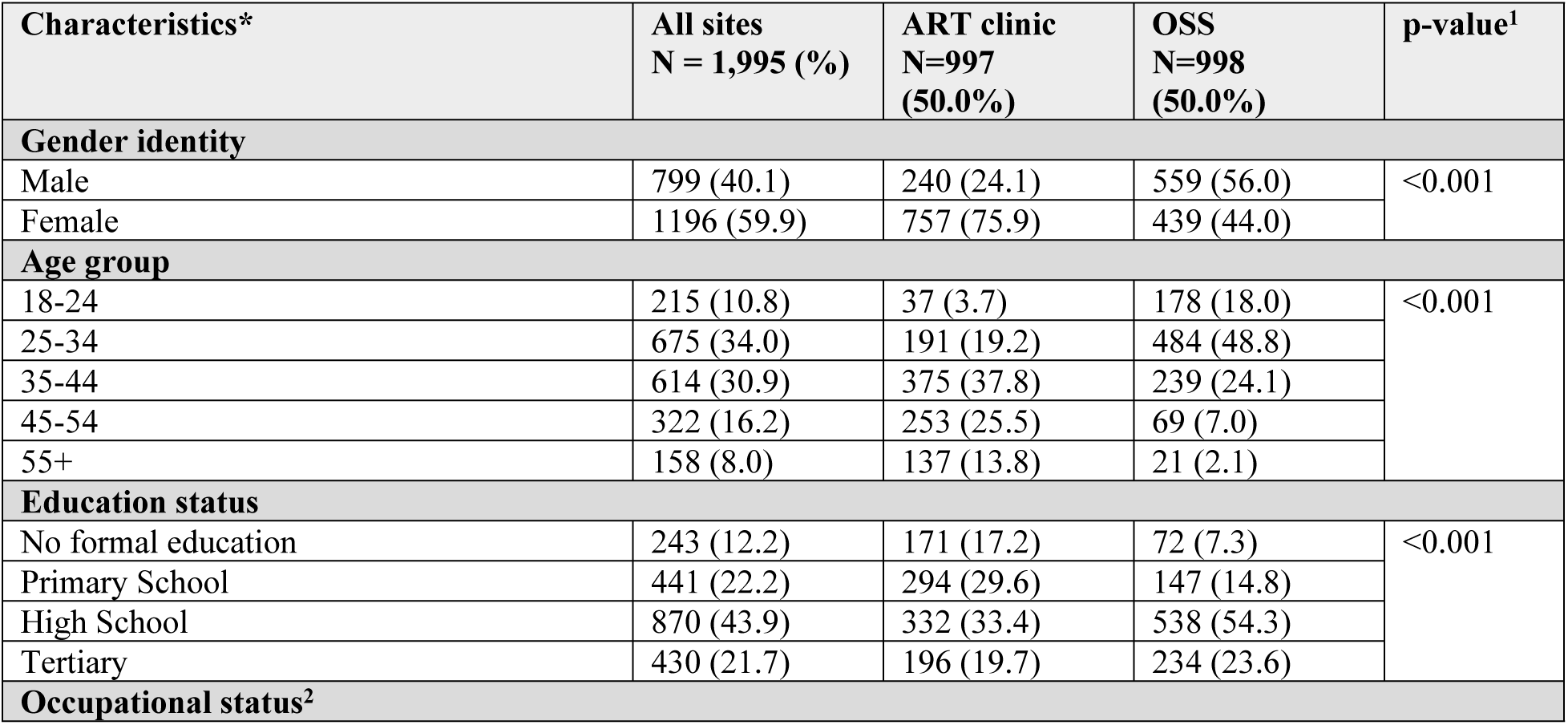
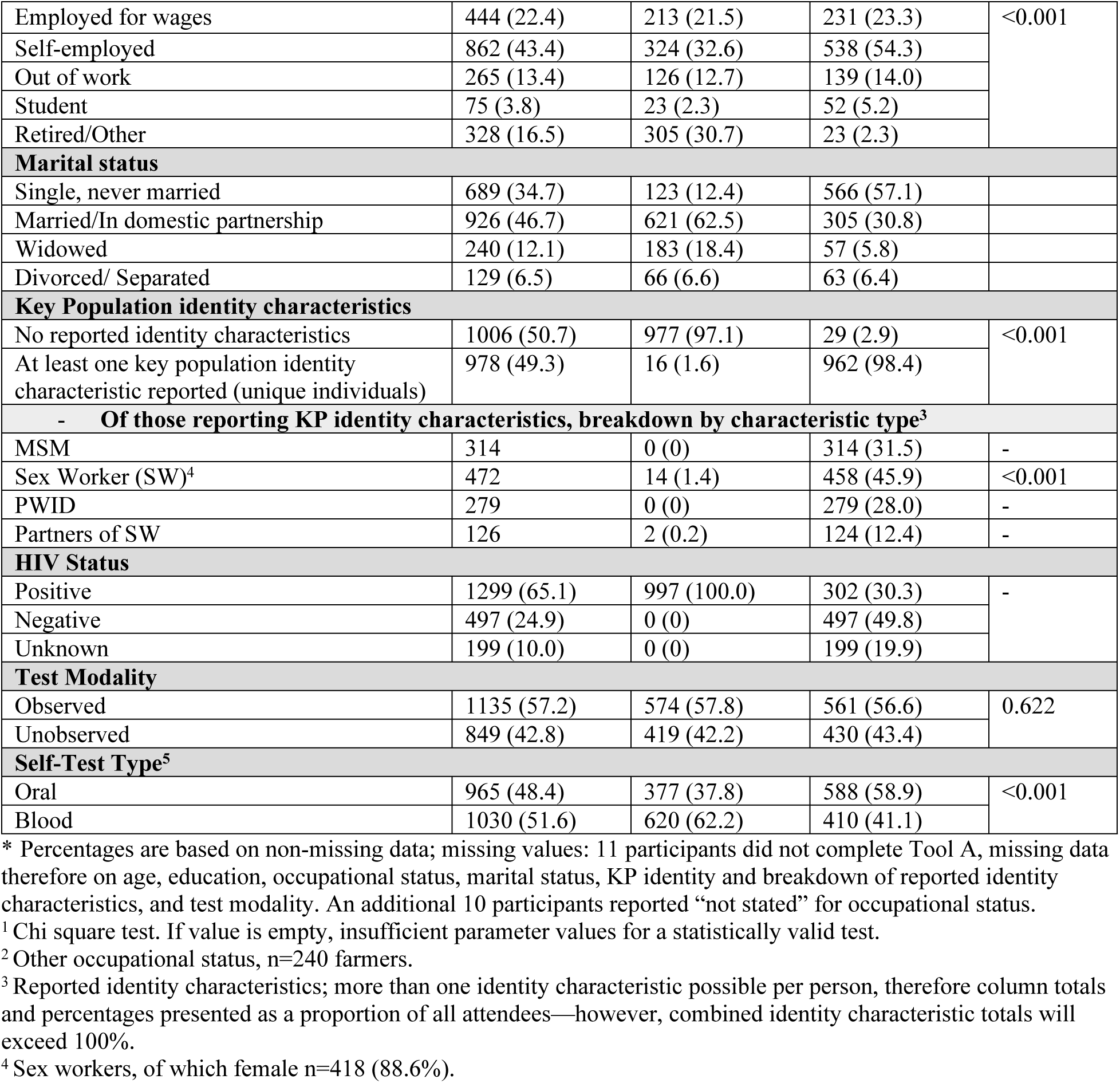
Participant characteristics.

Most participants used the observed testing modality (57.2%), indicating they used the HCVST on-site with assistance from a health worker, whereas 42.8% used the test either on-site or off-site with no assistance from a health worker. This was the case in both the ART clinic and the OSS sites. While the study aimed to distribute even numbers of blood-based and oral fluid HCVST across the four sites, more blood-based tests were distributed to participants than oral fluid tests (1,030 vs. 965, respectively). This was a result of the popularity of the blood-based kits—after 13 November 2023, clients could still enroll in the study but only had the option of oral-fluid tests. Observations about test type choice are thus descriptive only, not inferential. This lack of choice only affected the OSS (enrollment had finished in ART clinics), which likely contributes to the high proportion of clients in OSS using oral-fluid HCVST (58.9%), compared to ART clinics where the majority of clients used blood-based HCVST (62.2%).

### Feasibility

Feasibility was explored through study enrollment, test results and linkage across the clinical care cascade (**Figure 2**).

**Figure 2.**
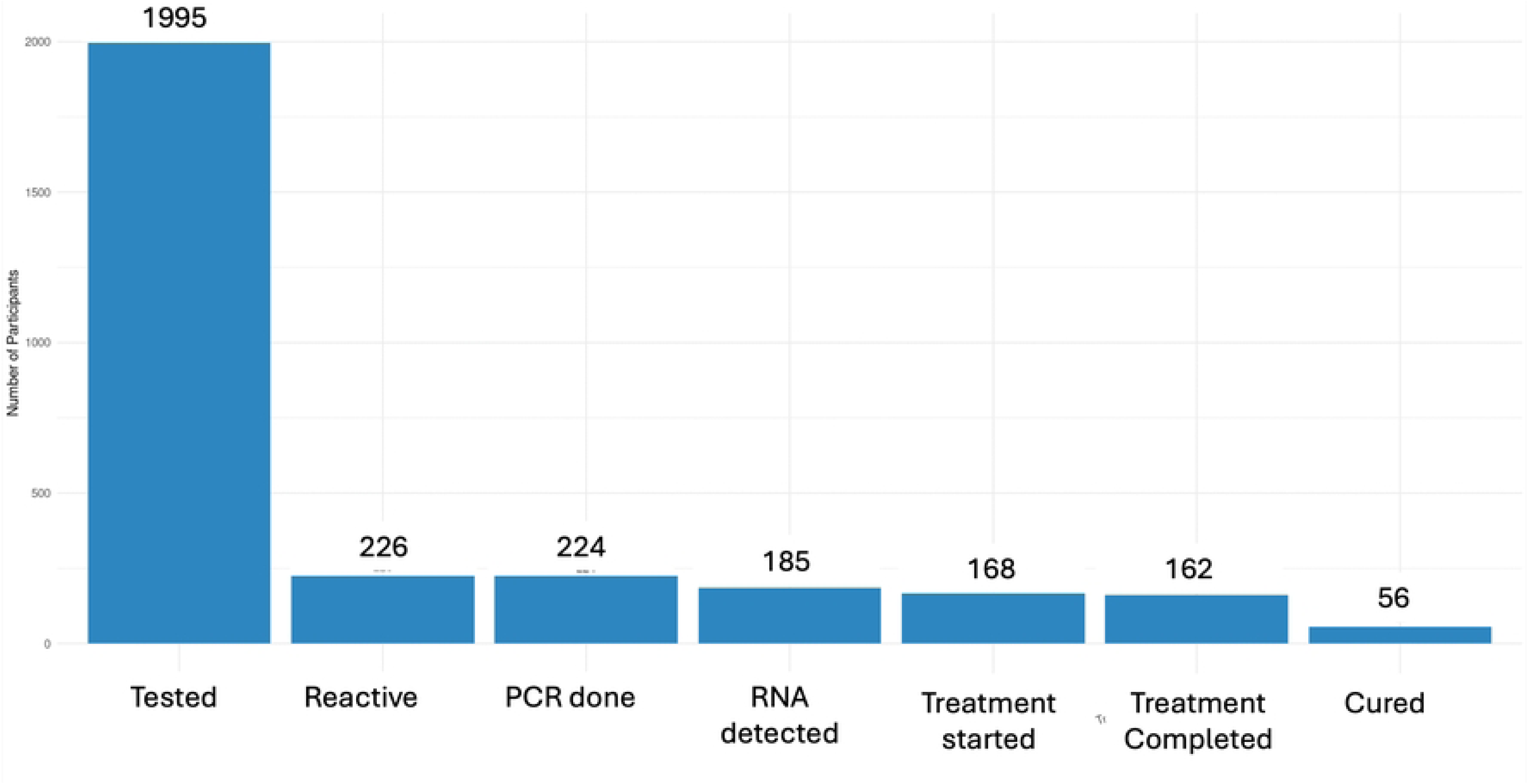
HCV clinical cascade care outcomes (absolute numbers)

Among 1,995 participants who used the HCVST, 226 (11.3%) had reactive test results. Of those, 224 (99.1%) received an RNA PCR test, and HCV RNA was detected in 185 (82.6%). Among persons with RNA detected, 168 initiated treatment, corresponding to a treatment initiation rate of 90.8%. Treatment completion was documented in 163 participants (97.0% of those initiating therapy), and sustained virologic response at 12 weeks post-treatment (SVR12, indicative of cure) was achieved in 56 participants, representing 34.4% of those who completed treatment. Differences were assessed by facility type (ART clinic vs. OSS; **Table 2**).

**Table 2.** HCV Clinical Cascade by Facility Type.

| Step | All Sites<br>N (%) | ART Clinic<br>N (%) | OSS<br>N (%) | p-value* |
| --- | --- | --- | --- | --- |
| Used HCVST | 1995 (100%) | 997 (100%) | 998 (100%) | - |
| HCVST reactive | 226 (11.3%) | 146 (14.6%) | 80 (8.0%) | <0.001 |
| RNA PCR performed | 224 (99.1%) | 144 (98.6%) | 80 (100%) | - |
| RNA detected | 185 (82.6%) | 115 (79.9%) | 70 (87.5%) | 0.207 |
| Treatment started | 168 (90.8%) | 110 (95.7%) | 58 (82.9%) | 0.008 |
| Treatment completed | 163 (97.0%) | 106 (96.4%) | 56 (96.6%) | - |
| SVR12 / Cured<sup>1</sup> | 56/61 (91.8%) | 36/36 (100%) | 20/25 (80.0%) | - |

\*Chi square test, values missing if any cell in the 2x2 table <5.
<sup>1</sup>Denominator for those achieving SVR12 includes only those participants who returned for viral load test and had available data at the end of the data collection period

Among individuals who used HCVST, the proportion with reactive test results was significantly higher at ART clinics compared to OSS (14.6% vs. 8.0%, *p* < 0.001). Among those with HCV RNA detected, treatment initiation was also higher at ART facilities, with 95.7% of RNA-positive individuals starting therapy compared to 82.9% at OSS facilities (*p*=0.008).

### Acceptability

Continuous acceptability scores were explored visually using histogram and boxplots by age, gender, educations status and facility type, with proportions in the lowest 10% of acceptability identified. HCVST was highly acceptable in this sample, with only a small minority indicating lower acceptability across empowerment, ease of use, and future use intentions (**Figure 3**).

**Figure 3.**
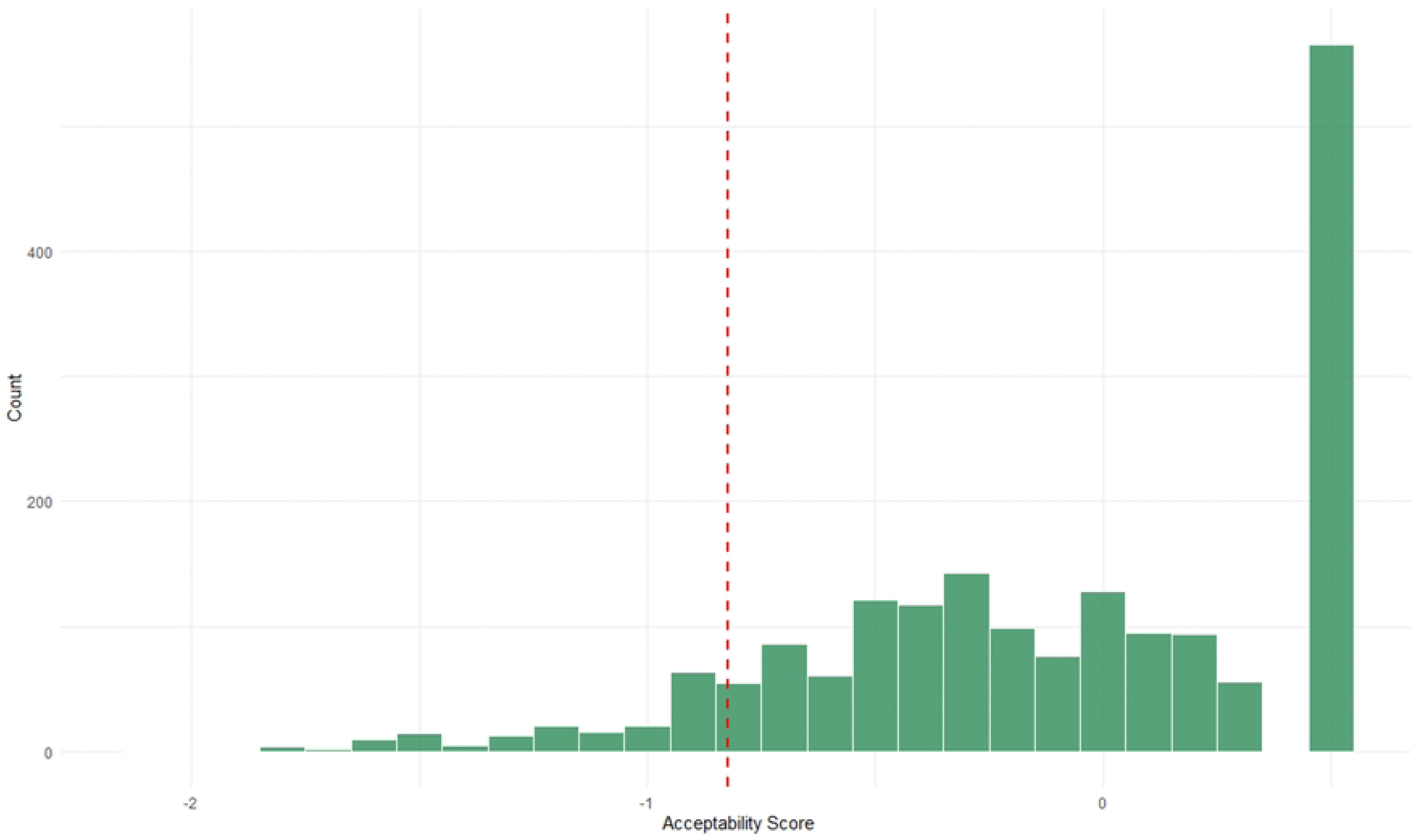
Distribution of acceptability score; values below the dotted red line indicate responses in the least 10% of acceptability.

Visible differences in distributions were observed by facility type, educational status and age, while differences by gender were minimal. OSS site attendance had lower acceptability scores, as did lowest educational status (no formal education) and younger age groups (18-24, and 25-34) had lower acceptability scores. The red cutoff line highlighted the proportion of participants falling within the lowest 10% of acceptability in each group (**Figure 4**).

**Figure 4.**
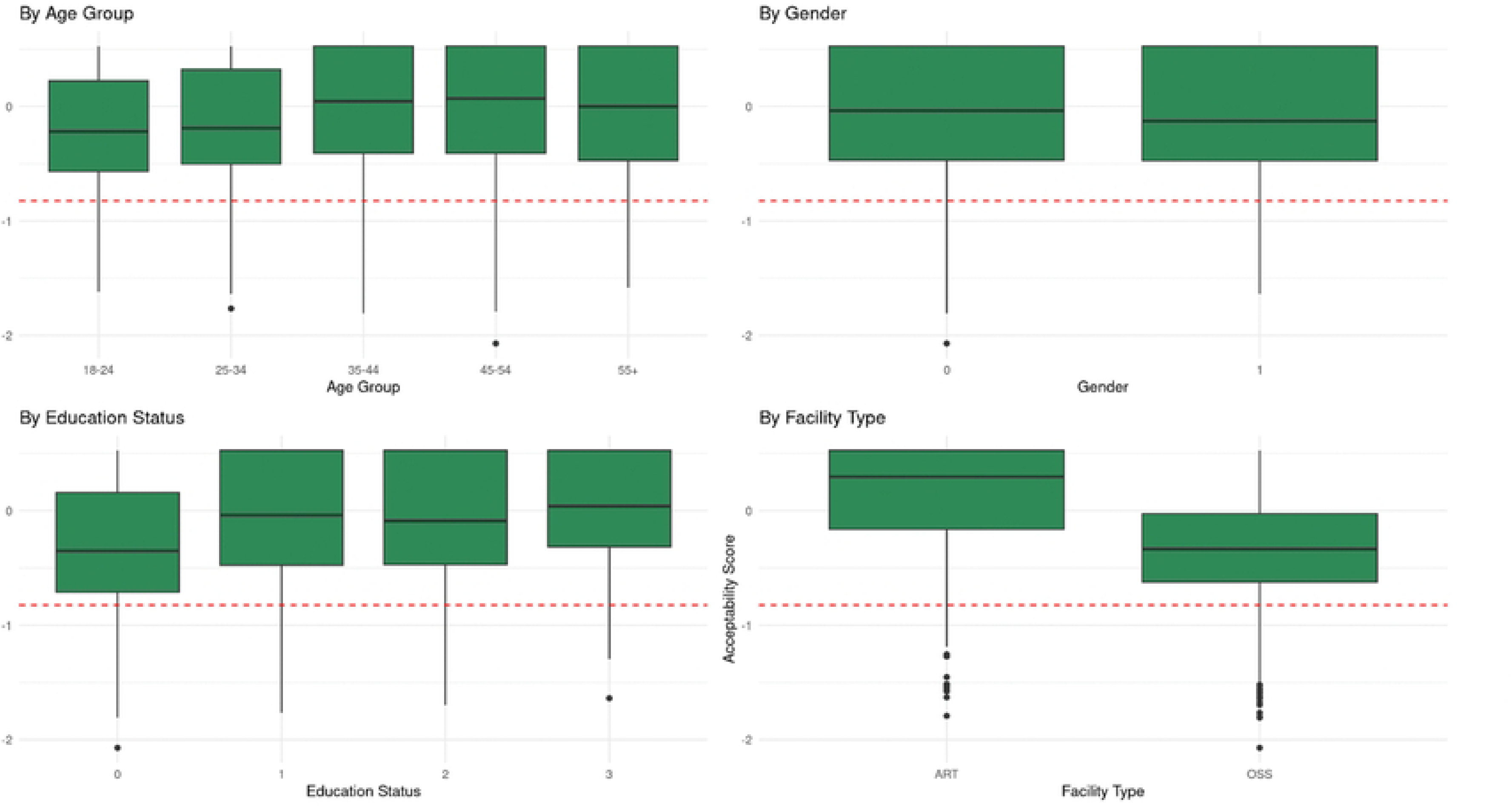
Box-and-Whisker plots of acceptability distribution by key covariates (age, gender, education, and facility type); Values below the dotted red line indicate responses in the least 10% of acceptability.

We modelled binary acceptability by facility type using logistic regression, adjusting for age, gender, and education status as prior confounders. Modelling facility type as the primary exposure in unadjusted analysis, participants at OSS sites had 2.3 times the odds of being in the lowest 10% of acceptability compared to those at ART clinics (OR = 2.3; 95% CI: 1.7–3.2; *p* < 0.001). After adjusting for age, sex, and education level, the association strengthened (aOR = 4.2; 95% CI: 2.8–6.2; *p* < 0.001) (**Table 4**).

**Table 4.** Odds of being in the lowest 10% of acceptability by Facility Type (N=1,868).

| Facility Type | Unadjusted OR<br>(95% CI) | Adjusted OR<br>(95% CI) | p-value* |
| --- | --- | --- | --- |
| OSS | 2.3 (1.7 to 3.2) | 4.2 (2.8 to 6.2) | <0.001 |
| ART Clinic | - | - |  |

\*Wald test

Qualitative analysis provided some potential explanatory detail both on the lower initiations of treatment at OSS and potentially lower acceptability. ART clients expressed a desire for more support when trying the self-tests but once trained, expressed very high confidence to use HCVST alone. They expressed a desire to “compare,” “be knowledgeable,” and “avoid mistakes,” suggesting a clear pathway from their initial need for assistance to independence. In contrast, participants in OSS were more likely to express a desire to use the HCVST alone, with some recognition that provider assistance was preferential, but only after attempting the test themselves. Attendees wanted to “just try something different”. As relates to feasibility, therefore, ART attendees appeared more likely to take the support offered in the clinic, whereas OSS attendees wanted to go it alone. This may reflect differences in clinic experience and engagement with formal health services on behalf of ART attendees that could be reflected in higher initiations on treatment once diagnosed.

However, acceptability appeared to be more associated with client characteristics than facility type: younger females (18-24 years) in the OSS expressed more concerns about needles and pain, as well as the highest initial need for guidance, and were less confident than their male counterparts. For example, “I will like to do it, in case I make a mistake, in the facility someone can guide me” (F, 18-24).

## Discussion

This study provides important insights into the feasibility and acceptability of HCVST among PLHIV and KP in Nasarawa State, Nigeria. HCVST was demonstrated as both feasible and acceptable in these populations. Feasibility was demonstrated by high uptake of services across the clinical cascade. Nearly all participants with reactive HCVST results accessed confirmatory RNA testing (99%), and of those with detected RNA, 90.8% started treatment. Treatment completion rates were also very high (97%), and for those who completed therapy and had a viral load test by the time the study ended, 92% achieved sustained virologic response (cure).

These outcomes indicate that HCVST can successfully prompt progression through the clinical cascade when accompanied by supportive linkage systems. HCVST was also highly acceptable overall among study participants.

Nevertheless, a critical observation in this study was the marked difference in treatment initiation and acceptability of the HCVST intervention based on facility type. While both ART clinics and OSS facilities demonstrated high engagement with HCVST and subsequent RNA PCR testing, treatment initiation rates were significantly higher in ART clinics compared to OSS (95.7% vs. 82.9%) and acceptability was also higher. This may suggest that established ART clinics that are already embedded in formal health systems may offer more effective patient navigation and support for linkage to care than OSS, or they may reflect unmeasured factors, such as staffing levels, on-site pharmacy support, or same-day drug availability. Patients in ART clinics are likely more clinically experienced than attendees of OSS, more used to formal health services, and more likely to accept clinician support to link into treatment; this is underlined by the findings of Sekhon et al., who posited that perceptions of acceptability of a healthcare intervention are influenced by participants’ and providers’ understanding of that intervention and how it works in relation to the problem it targets (“coherence”) [24]. Additional weight is given to this argument by the clear link from supportive testing intervention to independence amongst ART attendees in the qualitative results in this study. Although critical entry points for key populations, OSS may require strengthened referral and navigation systems to sustain downstream care, a finding with practical implications for program design and service integration.

This aligns with evidence from other LMIC contexts showing that decentralized or community-based facilities can successfully distribute self-tests but require additional attention for linkage to treatment [25–26]. Our findings are consistent with early studies on the introduction of HIV self-testing, which also showed generally positive findings and the need to tailor specific aspects—like linkage to treatment—for key populations in particular [27], and with a recent survey of Nigerians 18 years and above that showed generally strong support for HCVST among health workers and non-healthcare workers in Nigeria [28].

However, it is also important not to overstate these differences: differences in acceptability may reflect both true differences in perceptions or potential differences in how different participants across settings interpreted or responded to scale items.

Finally, we were surprised by the relatively low number of PWID taking up HCVST relative to other KP types from the OSS—low participation from this population type may also explain the lower reactivity among OSS clients. Nasarawa State has high HCV prevalence and strong outreach with waitlists for treatment, so it is possible that many PWID with known HCV status had already been reached.

### Implications

Overall, the high levels of feasibility and acceptability demonstrated in this evaluation, particularly for participants in ART clinics, support the broader introduction and scale up of HCVST in Nigeria as a means of expanding HCV diagnosis and treatment. HCVST integration in ART clinics may be particularly important for case finding and due to their successful linkage to treatment. However, targeted efforts are needed for OSS serving key populations and facilities with limited health system integration, including patient navigation, peer outreach, and stigma reduction.

Future research should aim to dissect the interrelated influences of key population identity, HIV status, and facility type, and further explore tailored interventions that address the unique needs and barriers faced by OSS users. Attention to qualitative insights will be crucial to inform strategies that enhance both feasibility and acceptability across diverse settings.

### Strengths and Limitations

Strengths of this study include the large sample size, a diverse mix of high-risk populations from ART clinics and OSS, smooth integration of HCVST into routine health services in the study sites, robust support for linkage to near point-of-care RNA PCR testing and free treatment, and the use of both quantitative and qualitative measures to assess feasibility and acceptability of HCVST.

The variability in sociodemographic characteristics and HCV reactivity across settings might reflect a selection bias in the study population, driven by the demographics of attendees at ART clinics and OSS. Given the high HCV prevalence in Nasarawa we expected to see more PWID among the KP at OSS in particular, and the low number of PWID may reflect a discomfort in revealing PWID identity even in KP-friendly spaces. Furthermore, although the same set of questions was used across all sites to identify KP identity characteristics, very few ART clinic participants reported KP identity characteristics. This is not unexpected, as health facilities generally have low rates of self-reported KP identity due to stigma and discrimination [29].

Allocation to blood- vs. oral-fluid HCVST was constrained by depletion of blood-based kits in November 2023, which coincided with slower enrolment at OSS. This introduced temporal and site confounding that limited our ability to compare acceptability by test type. Additionally, we did not assess whether the acceptability scale functioned equivalently across ART and OSS settings, so observed differences should be interpreted with that caveat. Lastly, our study was not designed to capture SVR12 outcomes for all participants—the SVR12 data reported in our care cascade are incomplete because follow up ended before the 12-week time frame passed for all participants.

## Conclusion

This study supports the expanding role of HCVST as a feasible and acceptable strategy to increase HCV diagnosis and linkage to care among high-risk populations in Nigeria, and advance the goals of Nigeria’s micro-elimination plans and broader elimination strategies. Policy makers and implementers should leverage the strengths of existing health services, including ART clinics and OSS, while ensuring flexible, stigma-sensitive approaches with robust support for linkage to RNA PCR testing and treatment.

## Data Availability

All relevant data are within the paper and its Supporting Information files.

